# Automated Estimation of Computed Tomography-Derived Left Ventricular Mass Using Sex-specific 12-Lead ECG-Based Temporal Convolutional Network

**DOI:** 10.1101/2024.02.19.24303061

**Authors:** Heng-Yu Pan, Benny Wei-Yun Hsu, Chun-Ti Chou, Chih-Kuo Lee, Wen-Jeng Lee, Tai-Ming Ko, Tzung-Dau Wang, Vincent S. Tseng

## Abstract

**Background:** Left ventricular hypertrophy (LVH) is characterized by increased left ventricular myocardial mass (LVM) and is associated with adverse cardiovascular outcomes. Traditional LVH diagnosis based on rule-based criteria using limited electrocardiogram (ECG) features lacks sensitivity. Accurate LVM evaluation requires imaging techniques such as magnetic resonance imaging or computed tomography (CT) and provides prognostic information beyond LVH. This study proposed a novel deep learning-based method, the eLVMass-Net, together with sex-specific and various processing procedures of 12-lead ECG, to estimate CT-derived LVM.

**Methods:** 1,459 ECG-LVM paired data were used in this research to develop a deep-learning model for LVM estimation, which adopted ECG signals, demographic information, QRS interval duration and absolute axis values as the input data. ECG signals were encoded by a temporal convolutional network (TCN) encoder, a deep neural network ideal for processing sequential data. The encoded ECG features were concatenated with non-waveform features for LVM prediction. To evaluate the performance of the predicting model, we utilized a 5-fold cross-validation approach with the evaluation metrics, mean absolute error (MAE) and mean absolute percentage error (MAPE).

**Results:** The eLVMass-Net has achieved an MAE of 14.33±0.71 and an MAPE of 12.90%±1.12%, with input of single heartbeat ECG waveform and lead-grouping. The above results surpassed the performance of best state-of-the-art method (MAE 19.51±0.82, P = 0.04; MAPE 17.62%±0.78%; P = 0.07) in 292(±1) test data under 5-fold cross-validation. Adding the information of QRS axis and duration did not significantly improve the model performance (MAE 14.33±0.71, P = 0.82; MAPE 12.90%±1.12%; P = 0.85). Sex-specific models achieved numerically lower MAPE for both males (−2.71%, P=0.48) and females (−2.95%, P=0.71), respectively. The saliency map showed that T wave in precordial leads and QRS complex in limb leads are important features with increasing LVM, with variations between sexes.

**Conclusions:** This study proposed a novel LVM estimation method, outperforming previous methods by emphasizing relevant heartbeat waveforms, inter-lead information, and non-ECG demographic features. Furthermore, the sex-specific model is a rational approach given the distinct habitus and features in saliency map between sexes.

**Clinical Perspectives:** *What is new?:*

- The eLVMass-Net used ECG encoders with lead grouping, a unique feature that more properly reflects the electrical orientation of left ventricle.
- The sex-specific deep learning model is able to discriminate inter-gender differences of ECG features as shown by saliency maps.

*What are the clinical implications?:*

- The eLVMass-Net outperforms current state-of-the-art deep learning models for estimating left ventricular mass.
- A more accurate estimation of left ventricular mass could improve quality of care for comorbidities such as hypertension from easily accessible ECG.

## Introduction

Left ventricular hypertrophy (LVH) is defined by an increased left ventricular myocardial mass (LVM), usually secondary to conditions with higher left ventricular afterload, such as hypertension or aortic stenosis. LVH is a dynamic pathophysiological phenomenon with concomitant changes in cardiomyocytes and interstitial fibrosis.^1,2^ Further progressions in LVH are associated with diastolic dysfunction, arrhythmia as well as cardiac death.^3–5^ Traditionally, the diagnosis of LVH relies on various rule-based criteria, mainly focusing on QRS voltage presented on individual electrocardiogram (ECG), which is a low-cost and convenient test.^6^ Nonetheless, most of these criteria concentrate more on the features of R/S amplitudes, QRS duration, and qualitative ST and T wave changes, which often fall short of sensitivity.^7–9^ It is not surprising considering the fact that magnetic resonance imaging (MRI)-based studies showed that commonly used criteria such as Sokolow-Lyon or Cornell indices are negatively correlated to the degree of left ventricular fibrosis.^10^ It is thus necessary that comprehensive ECG features, as well as interactions between individual leads, be considered in order to encompass the electrophysiological traits.^11^

More precise evaluation of LVH typically requires accurate imaging evaluation of LVM. Both cardiac MRI and computed tomography (CT) are recommended as accurate measurement modalities of LVM and are able to provide additional prognostic value beyond LVH.^12,13^ However, these imaging modalities are either time-consuming or flawed by radiation and contrast exposure. Also, such information provides anatomical rather than electrophysiological features.

In the past, machine learning (ML) models have been applied to ECG features generated through rule-based algorithms, but these methods have limitations in producing high-quality ECG features. To address this issue, some works employed learning-based techniques, specifically deep learning models, to replace rule-based algorithms in ECG feature extraction. A few studies have made progress in LVM evaluation using ECG amplitude data or auto-segmented features.^14–16^ Some studies take advantage of convolutional neural networks for heart disease (e.g., LVH) prediction.^17,18^ The ecgAI model used a deep-learning model to automatically segment raw ECG signals into non-overlapping intervals and durations to generate ECG features.^14^ This approach allowed for a more comprehensive analysis of the ECG signal, resulting in more accurate and reliable features. Furthermore, the LVM-AI model utilized an end-to-end training pipeline to estimate LVM using ECG signals and demographic data.^16^ By incorporating demographic data in the analysis, they were able to improve the accuracy of LVM prediction. These deep learning methods that discriminate LVH may improve risk stratification and prompt early pharmacological intervention.^19^

Previous studies utilized full-length ECG signals and demographic data as the input for their models. However, it is difficult to ensure that the model is able to focus specifically on the waveform features that are related to LVM values or to extract inter-lead information such as the heart axis. In this study, to solve this problem, we developed a deep-learning model for LVM estimation based on the Taiwan CVAI dataset, which includes cardiac CT exams of over 3,500 patients from major medical centers in Taiwan. We accessed demographic data, 12-lead ECG, and CT-derived LVM values, utilizing these data to construct an LVM estimation model. Meanwhile, we conducted a series of experiments to analyze the impact of sex, ECG preprocessing methods, and groupings for 12-lead ECGs according to different characteristics of model performance.

## Methods

### Data Acquisition

The dataset utilized in this study was obtained from the National Taiwan University Hospital and was approved by the institutional review board previously. This study was further approved by the Institutional Review Board of National Taiwan University Hospital (NTUH-REC No. 202012128RINA). The dataset consists of 12-lead ECG signals, recorded at a sampling rate of 500 Hz for 10 seconds and downloaded in XML format. The XML files also contain non-waveform information automatically generated by the ECG device, including heart axis and QRS duration, which are further integrated into input features. Pertinent demographic data such as age, sex, height, and weight are included. **Figure 1** shows the overall data collection and cleansing process. The LVM values were first obtained, and subsequently, the ECG XML file that was closest to the LVM measurement was selected as the corresponding ECG signal. Patients with bundle branch block, paced rhythm, or atrial fibrillation were excluded from the research.

**Figure 1.**
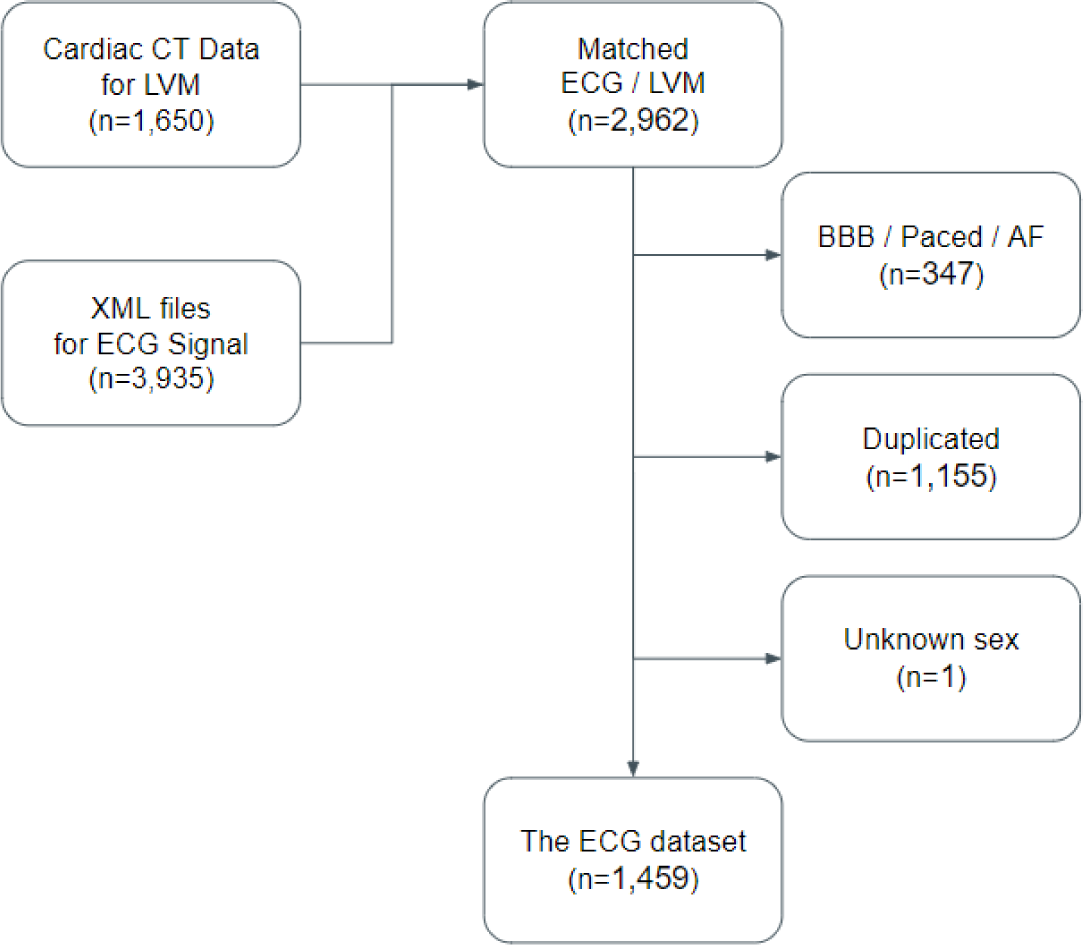
Overview of the dataset collection. The CT data and corresponding XML files were collected independently. Therefore, a matching process was carried out based on the requirement that both measurements be taken within six months. Patients with bundle branch block (BBB), paced rhythm, and atrial fibrillation (AF) were excluded due to distorted ECG waveforms. Additionally, ECG recordings that did not have a one-to-one paired LVM measurement were also excluded. As a result, a total of 1,459 valid data points were included in this study.

The ground truth of LVM values was inferred from auto-segmentation of left ventricular wall from cardiac CT images, by using the Intellispace Portal Software (Philips Healthcare, The Netherlands). A threshold-based method was used to determine epicardial and endocardial borders, and the left ventricular myocardium was calculated automatically after obtaining both total ventricular volume and ventricular cavity volume. Mass value was further acquired after multiplying myocardial density by 1.05 g/mL. The results of left ventricle segmentation were verified by a senior radiologist (W.-J.L.) who is specialized in cardiac CT images and with more than 20 years’ experience.

The dataset is divided into non-overlapping subsets for cross-validation. K-fold cross-validation was employed as a robust validation process to prevent sampling bias. In each iteration, a single fold was reserved as the test set (n=292±1), while a percentage of the training set (n=1051±1) was selected as the validation set (n=116±1) to assess model performance. The selection of the optimal model was based on the performance observed during the validation stage. The final evaluation of the model was obtained by averaging the test results across all folds. All experiments in this research undergo validation using 5-fold cross-validation, which is stratified based on the LVM values. For sex-specific models, the original data splitting policy was followed, and the samples of the target gender were extracted to form the sub-datasets for training and evaluation.

### Data Processing

We included demographic information (age, sex, height, and weight), and automatically derived numeric ECG values (P-axis, R-axis, T-axis, and QRS duration) for analysis. These data were represented as scalar values, resulting in a total of 8 scalar inputs for this task. Our research aimed to leverage both the ECG signals and non-ECG data to estimate the corresponding LVM value. cr data were addressed through an imputation method, with numerical data imputed with the median and binary categorical data with 0.5.

The ECG signals were in the form of a 12-lead signal with a shape of (L, D * Fs), where L represents the number of leads, D represents the signal duration in seconds, and Fs represents the sampling rate. This study improved the quality of 12-lead ECG signals through various preprocessing steps. The signals underwent a band-pass filter to eliminate high-frequency noise and baseline wandering. R peaks were then detected, and the middle heartbeat segment was selected to avoid incomplete segments caused by recording borderlines.

### eLVMass-Net

This study has developed a novel deep learning-based method, the eLVMass-Net, which can accurately estimate LVM values using ECG and demographic information. The overview of the proposed framework is depicted in **Figure 2**. There are ECG feature extractors (i.e., the encoders for input data embedding) followed by a multilayer perceptron (MLP) layer. The number of ECG encoders utilized in the model is dependent on the number of lead groups. The encoded ECG features from each ECG encoder were concatenated, and a projection layer was utilized to aggregate these features. The scalar features such as demographic data, axis, and QRS duration were passed through their own MLP layer. These two feature vectors were subsequently concatenated and fed into an MLP regressor to estimate the LVM value. To obtain ECG features, we took advantage of the Temporal Convolutional Networks (TCN) to encode ECG signals. Throughout the process of model training, the mean absolute error (MAE) was employed as the chosen loss function. The Adam optimizer, with a learning rate of 0.001, was utilized for model optimization. And with a maximum of 100 epochs, the model with the lowest validation loss was selected for testing to avoid overfitting.

**Figure 2.**
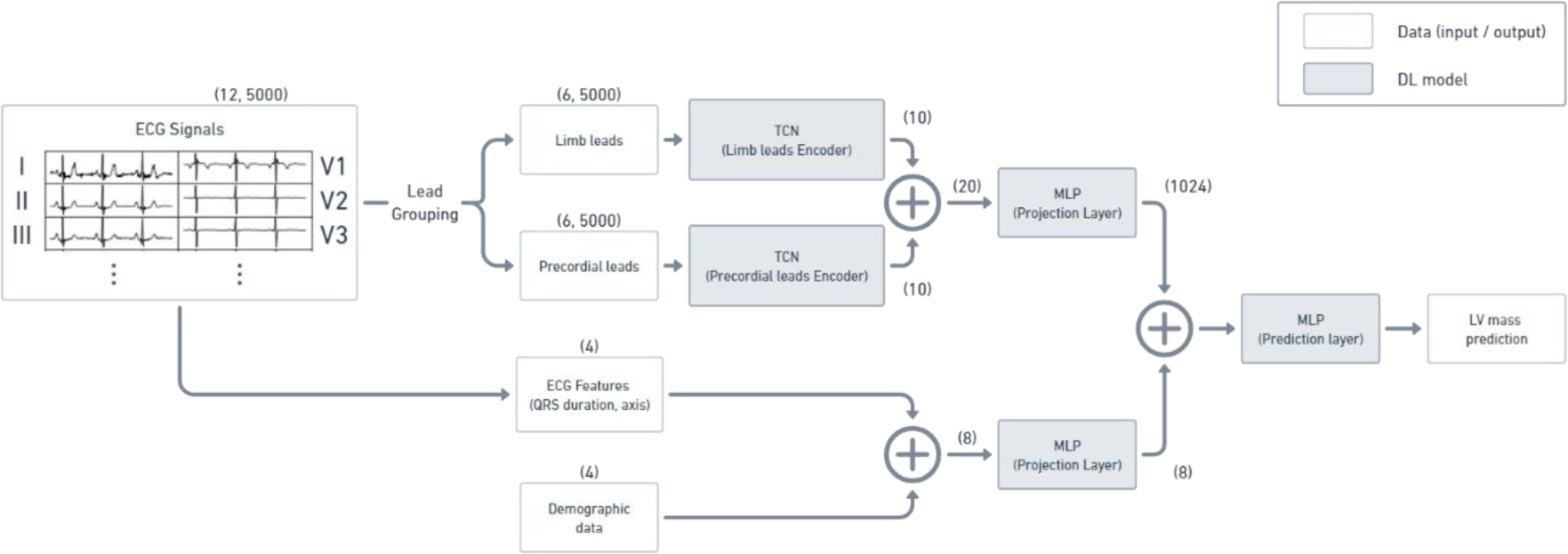
Overview of the proposed LVM estimation framework. The proposed LVM prediction model consists of separate encoders for the limb leads and chest leads of the 12-lead ECG, followed by a multilayer perceptron (MLP) layer. Additionally, scalar features such as demographic data, heart axis, and QRS duration are passed through their own MLP layer. The encoded ECG features and scalar features are then concatenated and fed into the prediction layer to estimate the LVM.

Besides TCN, we also used EfficientNet for comparison to validate the performance of the convolutional neural network (CNN)-based methods with different characteristics. EfficientNet and TCN are two popular models used for image and signal processing. EfficientNet is proposed based on a CNN architecture, which has demonstrated exceptional performance in image classification tasks and even on ECG signals.^20–22^ These models are designed using a compound scaling method that optimizes the network’s depth, width, and resolution. The EfficientNet architecture also employs advanced techniques like squeeze-and-excitation modules and swish activations, which further enhance its performance. Meanwhile, TCN is a type of deep neural network that is ideal for processing sequential data, such as time-series signals. TCNs use dilated convolutions, enabling the network to capture long-term dependencies in the input sequence while maintaining a compact architecture. Our preliminary experiments revealed that EfficientNet-based ECG encoders had a tendency towards overfitting.

### Exploring ECG Pre-processing and Grouping Methods

We explored the use of preprocessing techniques to guide the model to learn ECG features. We employed two preprocessing techniques, namely random length crop and single heartbeat extraction. The random length crop approach involves randomly selecting a segment of the ECG signal of varying lengths and using it as input to the model. This approach enables the model to learn features that are specific to different parts of the ECG waveform, which may be useful in capturing subtle changes in the signal. On the other hand, the single heartbeat extraction approach involves segmenting the ECG signal into individual heartbeats and using each beat as input to the model. This approach may help the model to focus on capturing features that are specific to the heartbeat waveform, instead of inter-beat waveform variances. Furthermore, we investigated the use of lead grouping as a preprocessing technique for improving the prediction accuracy of ECG signals. Specifically, we applied lead grouping on the 12-lead ECG signals based on their electrical orientation or anatomical location. For electrical orientation, 12-lead ECG signals are grouped into 2 separate groups based on horizontal and frontal planes (i.e., precordial and limb leads). For anatomical location, we formed 4 groups based on the distribution of coronary artery branches within the heart. Leads V1-V4 were grouped as leads related to the left anterior descending artery. Leads I, aVL, V5, and V6 were grouped as leads related to the left circumflex artery, whereas leads II, III, and aVF and lead aVR were related to the right coronary artery and the left main coronary artery, respectively.

### Feature Importance

To better understand how different input data contribute to the overall prediction performance of our model, we designed three input combinations in our study. The first input set included only raw ECG signals, which is a commonly used and simple setting that can be applied to any 12-Lead ECG device and dataset. This setting served as a baseline for comparison with more complex input sets. Demographic data was added to the second input set, as this information can provide a general description of physiological conditions that may affect electric conductance from the heart and heart functions. Demographic data are widely available in most clinical fields, making it a useful addition to the input set. Lastly, we included ECG axis and QRS duration information extracted from the XML files of ECG devices. However, this input combination may not always be available, as not all ECG devices provide this information.

By comparing the performance of the three input combinations, we were able to identify the contributions of each input data type to the overall prediction performance of our model. This allowed us to determine which input combinations were most effective for predicting LVM.

### Performance Analysis

To evaluate the performance of the predicting model, mean absolute error (MAE) and mean absolute percentage error (MAPE) were utilized. These widely accepted metrics are commonly employed in regression analysis to gauge the accuracy of predicted values in comparison to actual values. MAPE calculates the absolute percentage difference between predicted and actual values and averages them over the dataset, while MAE measures the average absolute difference between the predicted and actual values.

We also used the saliency maps from the proposed framework to assess the importance of different ECG segments. Saliency maps are a type of visualization tool that can be generated from deep learning models to help understand how the model is making its predictions. These maps highlight the most important regions of input data that the model focuses on when making its prediction. In our case, saliency maps can be used to visualize which parts of the ECG signal are most important for the model’s prediction. For the illustration of saliency maps, the ECG signal was divided into non-overlapping segments, including the PR interval, QRS interval, ST segment, T-wave, and TP interval (**Figure S1**). Subsequently, the significance of each segment was determined by calculating the summation of their respective significance scores, which represent the overall segment significance.

## Results

### Patient Characteristics

The sex-specific patient characteristics are summarized in **Table 1**. Compared with female patients, male patients generally had wider QRS segments and more leftward axis, while also having higher LVM and LVM index. The correlation between LVM values and demographics, QRS duration, or axis is shown in **Figure S2**.

**Table 1.** Characteristics for the dataset.

|  | **Male (n = 940)** | **Female (n = 519)** | **P value** |
| --- | --- | --- | --- |
| LV mass (g) | 123.29 (±29.88) | 85.21 (±21.41) | <0.01 |
| LV mass index (g/m<sup>2</sup>) | 67.43 (±14.71) | 52.98 (±13.02) | <0.01 |
| Age | 60.29 (±11.29) | 62.99 (±11.15) | <0.01 |
| Height | 168.59 (±6.12) | 158.16 (±6.39) | <0.01 |
| Weight | 72.86 (±11.53) | 60.47 (±9.36) | <0.01 |
| QRS-Duration | 93.86 (±10.74) | 86.26 (±9.63) | <0.01 |
| P-Axis | 49.52 (±20.76) | 49.85 (±23.30) | 0.78 |
| R-Axis | 28.45 (±35.37) | 36.79 (±34.21) | <0.01 |
| T-Axis | 40.29 (±32.98) | 43.47 (±33.05) | 0.08 |

### Effect of ECG Grouping and Preprocessing Methods

For the assessment of ECG preprocessing, data from both sexes were utilized as the training data, and electric orientation-based grouping was employed, whereas single-heartbeat extraction was utilized as the pre-processing technique for the assessment of different lead grouping methods.

The results of using different ECG preprocessing techniques are shown in **Table S1**. We found that the synchronized single heartbeat extraction can better improve the performance of the deep learning model in predicting LVM. Meanwhile, the experiments on lead grouping suggested that grouping based on electrical orientation had a significant impact on the performance of the model (**Table S2**).

### Feature Importance

The results of different input combinations are shown in **Table 2**. Demographic data played a crucial role in predicting LVM using our model. The inclusion of demographic data significantly improved the prediction performance compared to using only raw ECG signals by 25.1% (P<0.01). Furthermore, the addition of the heart axis and QRS duration information provided an insignificant performance improvement (by an absolute difference of 0.7%, P=0.82). In the real clinical setting, while demographic data are typically accessible, the availability of heart axis and QRS duration relies on the specific ECG device being used. Therefore, in the following experiments, both models were compared. The first model, named eLVMass-Net model 1, is trained using ECG and demographic data within the proposed framework. On the other hand, eLVMass-Net model 2 represents the proposed model trained with ECG, demographic data, heart axis, and QRS duration, encompassing a more comprehensive set of input features.

**Table 2.** LVM prediction performance using different input combinations. The relative improvements were computed by comparing the results obtained from the multimodal models with those of the ECG-only model. Furthermore, the P-values were calculated by comparing these methods to the outcomes of eLVMass-Net model 2.

| Feature Combinations | MAE |  | P-Value | MAPE |  | P-Value |
| --- | --- | --- | --- | --- | --- | --- |
|  | MAE | Relative Improvement |  | MAPE | Relative Improvement |  |
| ECG | 19.22<br>($\pm 0.91$) | - | <0.01 | 17.39%<br>($\pm 1.39\%$) | - | 0.02 |
| ECG + Demographics<br>(eLVMass-Net model 1) | 14.56<br>($\pm 0.53$) | 24.2% | 0.82 | 13.03%<br>($\pm 1.00\%$) | 25.1% | 0.85 |
| ECG + Demographics + Axis<br>(eLVMass-Net model 2) | 14.33<br>($\pm 0.71$) | 25.4% | | 12.90%<br>($\pm 1.12\%$) | 25.8% | - |

### Comparison with The State-of-The-Art Methods and Sex-Specific Analysis

We conducted performance comparisons using two different feature sets. The first setting followed the original configuration of the state-of-the-art (SOTA) methods, which involved specific method designs that were incompatible with our proposed eLVMass-Net. And in the second setting, all available features (ECG signals, demographic data, heart axis, and QRS durations) are used, showcasing the advantages of our approach while improving the SOTA methods as well.

For the non-sex-specific models, the performance metrics of our proposed method and the other SOTA methods are summarized in **Table 3**. As can be seen from the table, the proposed method has achieved the lowest mean absolute error (MAE) of 14.33 and mean absolute percentage error (MAPE) of 12.90% among all the methods. On the other hand, the SOTA methods have MAE and MAPE of 19.51 and 17.62%, respectively.

**Table 3.** The performance of the proposed prediction module and the SOTA models (testing sample n = 292±1). The P-values were computed by comparing these methods to the results of eLVMass-Net model 2.

|  | MAE |  | P-Value | MAPE |  | P-Value |
| --- | --- | --- | --- | --- | --- | --- |
| Model Name | Original Setting | All Features |  | Original Setting | All Features |  |
| ecgAI | 29.62<br>($\pm 0.93$) | 21.28<br>($\pm 0.36$) | <0.01 | 26.85%<br>($\pm 1.15\%$) | 19.12%<br>($\pm 0.56\%$) | <0.01 |
| LVM-AI | 19.58<br>($\pm 0.94$) | 19.51<br>($\pm 0.82$) | 0.04 | 17.52%<br>($\pm 1.25\%$) | 17.62%<br>($\pm 0.78\%$) | 0.07 |
| eLVMass-Net model 1<br>(ECG + Demographics) | - | 14.56<br>($\pm 0.53$) | 0.82 | - | 13.03%<br>($\pm 1.00\%$) | 0.85 |
| eLVMass-Net model 2<br>(ECG + Demographics + Axis) | - | 14.33<br>($\pm 0.71$) | | - | 12.90%<br>($\pm 1.12\%$) | - |

In the sex-specific analysis, each sex-specific model was trained and evaluated on sex-specific data subsets. The performance metrics for the sex-specific models with the proposed method and the other SOTA methods are summarized in **Table 4**. It’s observed that all methods are able to achieve a lower MAPE on the sex-specific dataset. Both eLVMass-Net Model 1 and eLVMass-Net Model 2 outperform the SOTA method (LVM-AI) by at least 3.68% (P < 0.01) and 2.21% (P = 0.20) in terms of MAPE for males and females, respectively. The results also indicated that models tend to have higher MAE for males and lower MAE for females compared to the MAE for the overall test set. This observation can be attributed to the higher average LVM value of males compared to females. When compared to the non-sex-specific model, the sex-specific model demonstrates a relative improvement of 2.71% in terms of MAPE for males (P = 0.30), and a relative improvement of 2.95% for females (P = 0.10). (**Table S3**)

**Table 4.**
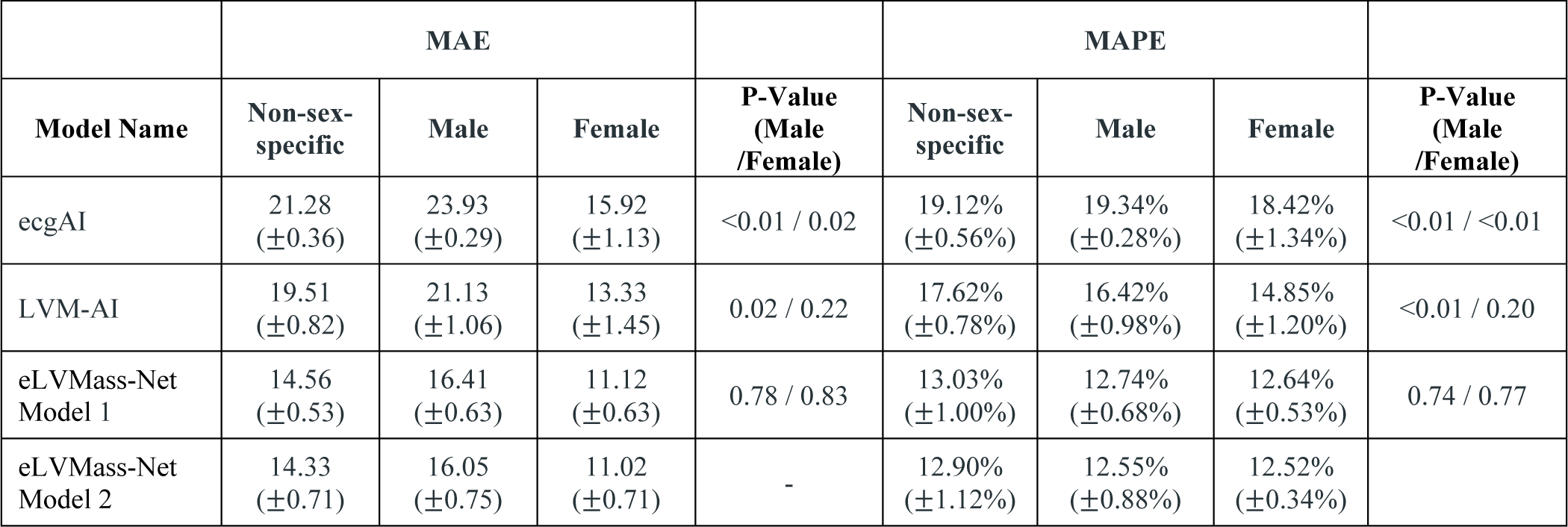
Sex-specific model performances of proposed prediction module and SOTA models (testing sample n = 292±1). The P-values were computed by comparing these methods to the results of eLVMass-Net model 2.

|  | MAE |  |  |  | MAPE |  |  |  |
| --- | --- | --- | --- | --- | --- | --- | --- | --- |
| Model Name | Non-sex-specific | Male | Female | P-Value (Male /Female) | Non-sex-specific | Male | Female | P-Value (Male /Female) |
| ecgAI | 21.28<br>($\pm 0.36$) | 23.93<br>($\pm 0.29$) | 15.92<br>($\pm 1.13$) | <0.01 / 0.02 | 19.12%<br>($\pm 0.56\%$) | 19.34%<br>($\pm 0.28\%$) | 18.42%<br>($\pm 1.34\%$) | <0.01 / <0.01 |
| LVM-AI | 19.51<br>($\pm 0.82$) | 21.13<br>($\pm 1.06$) | 13.33<br>($\pm 1.45$) | 0.02 / 0.22 | 17.62%<br>($\pm 0.78\%$) | 16.42%<br>($\pm 0.98\%$) | 14.85%<br>($\pm 1.20\%$) | <0.01 / 0.20 |
| eLVMass-Net Model 1 | 14.56<br>($\pm 0.53$) | 16.41<br>($\pm 0.63$) | 11.12<br>($\pm 0.63$) | 0.78 / 0.83 | 13.03%<br>($\pm 1.00\%$) | 12.74%<br>($\pm 0.68\%$) | 12.64%<br>($\pm 0.53\%$) | 0.74 / 0.77 |
| eLVMass-Net Model 2 | 14.33<br>($\pm 0.71$) | 16.05<br>($\pm 0.75$) | 11.02<br>($\pm 0.71$) | - | 12.90%<br>($\pm 1.12\%$) | 12.55%<br>($\pm 0.88\%$) | 12.52%<br>($\pm 0.34\%$) | |

Samples (n=5 for each) of low, medium, and high LVM values were selected. The mid part of the T wave in precordial leads and the QRS segment in limb leads are highlighted as important features with increasing LVM. **Table 5** presents a non-sex-specific summary of the segment-wise significance of the input ECG. It shows that the importance is mainly concentrated in QRS interval and T-wave. Furthermore, the importance of precordial leads decreased as the LV mass value increased (62.16% for the low LVM group and 40.02% for the high LVM group).

**Table 5.**
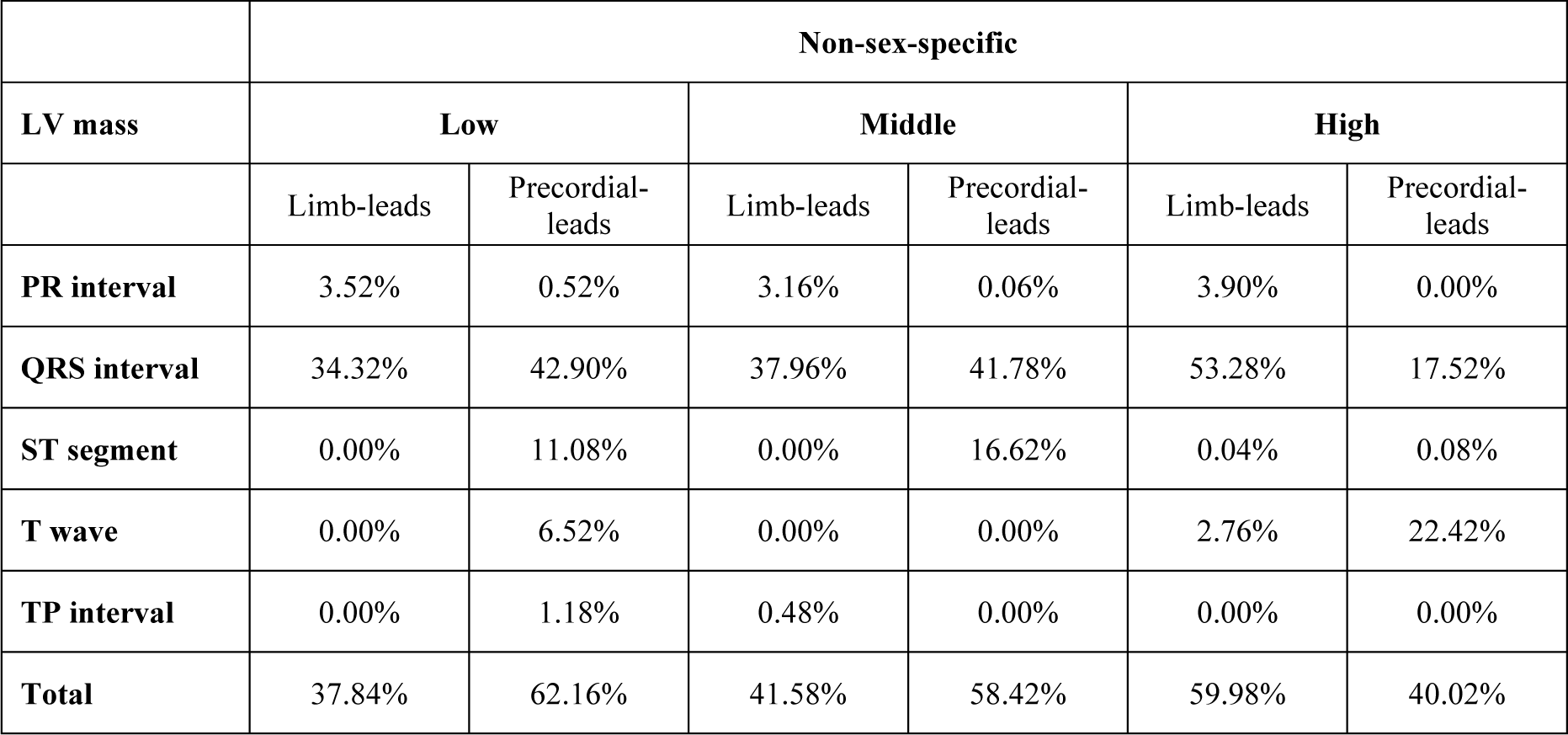
ECG-segment-wise importance for each segment from saliency maps from the non-sex-specific model. The percentages were the averages from 5 samples.

The segment-wise importance for sex-specific model is shown separately in **Table 6**. The saliency map for sex-specific model is illustrated in **Figure 3**. Notably, the importance of T waves experiences a proportional augmentation for males with increasing LVM values (3.36% for low LVM to 25.44% for high LVM). This trend has not been observed in females (0.84% in low LVM to 0.66% in high LVM). Conversely, female presented with persistently higher significance of the precordial QRS segment (62.26% in low LVM to 54.32% in high LVM).

**Figure 3.**
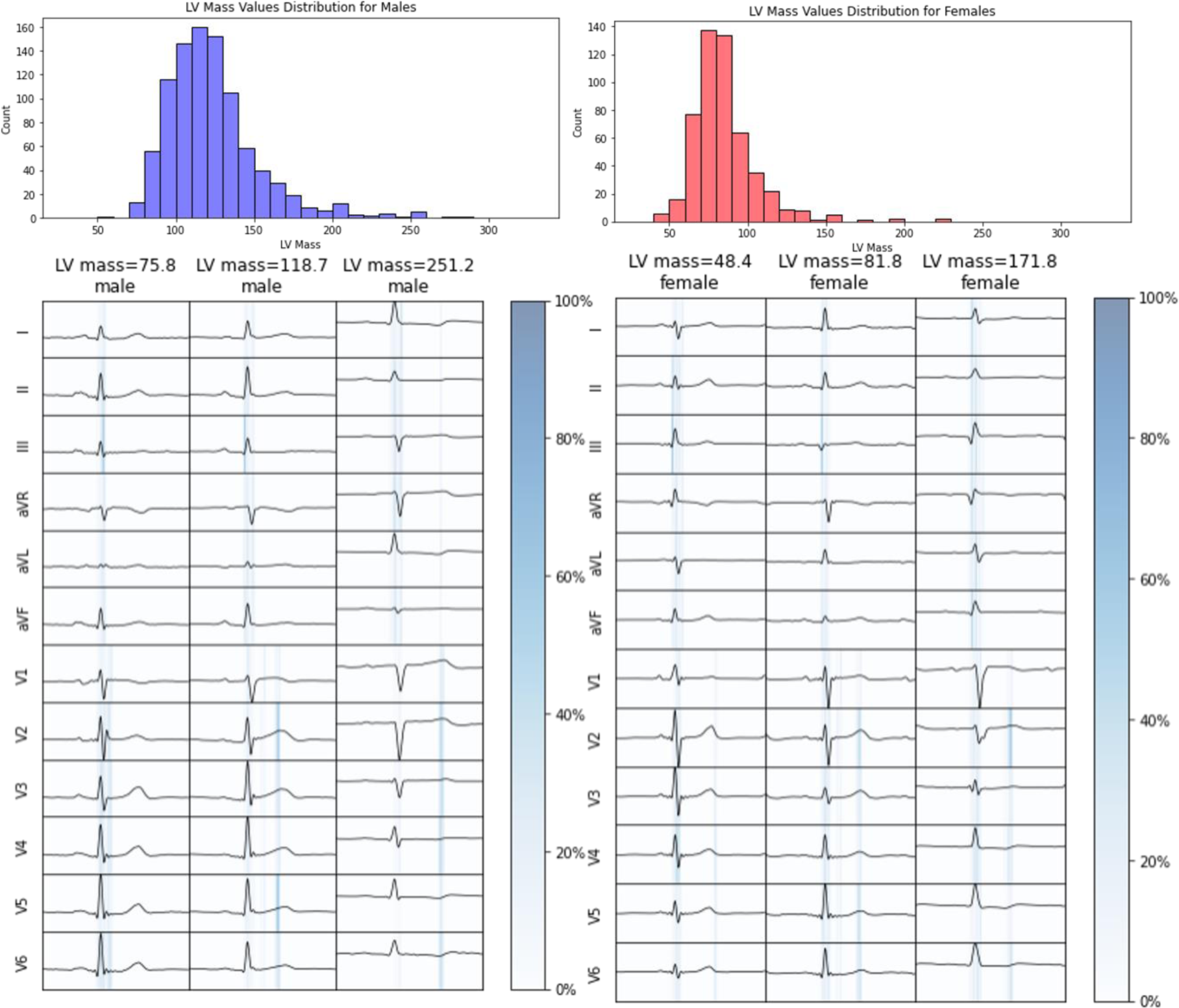
Saliency maps of the proposed prediction module. Three samples were selected to represent low, middle, and high LVM values for both males and females.

**Table 6.**
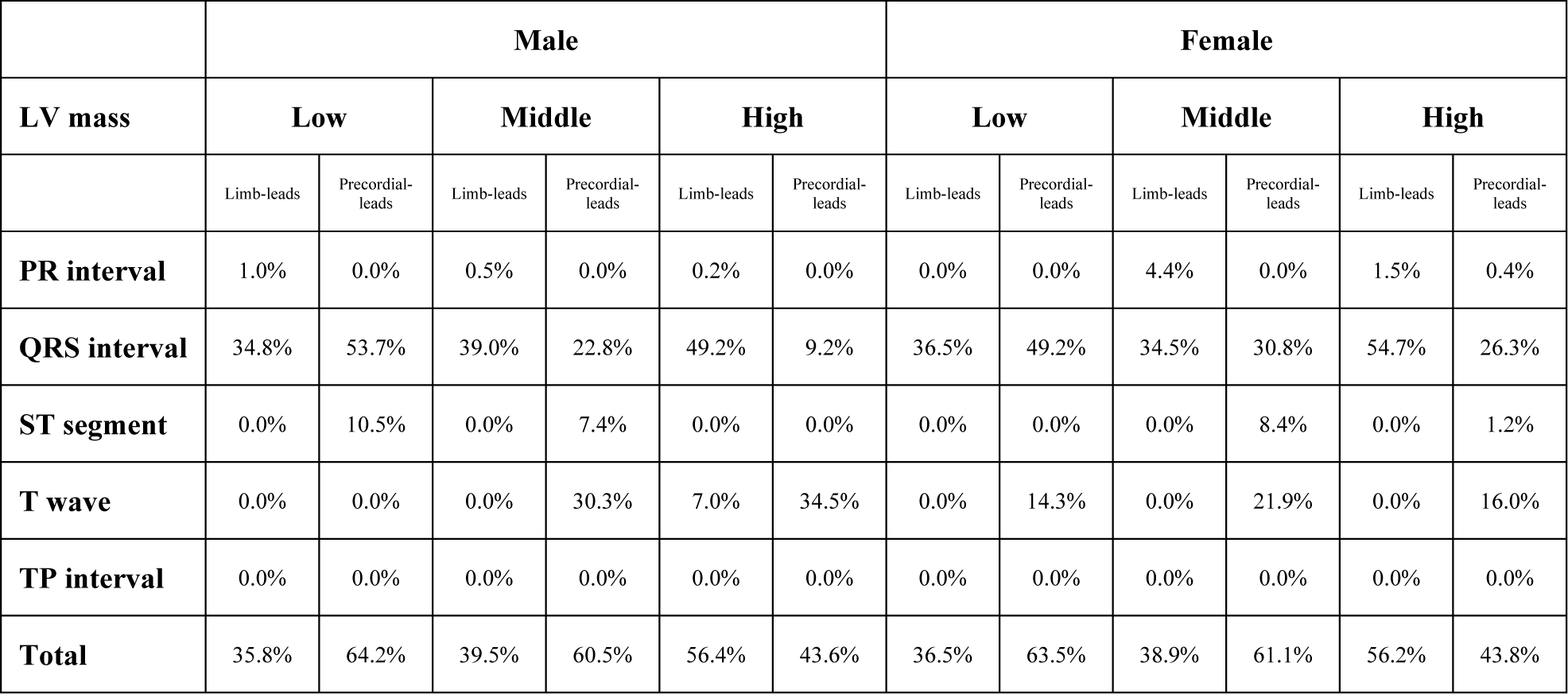
ECG-segment-wise importance for each segment from saliency maps from the sex-specific models. The percentages were the averages from 5 samples.

|  | Male |  |  |  |  |  | Female |  |  |  |  |  |
| --- | --- | --- | --- | --- | --- | --- | --- | --- | --- | --- | --- | --- |
| LV mass | Low |  | Middle |  | High |  | Low |  | Middle |  | High |  |
|  | Limb-leads | Precordial-leads | Limb-leads | Precordial-leads | Limb-leads | Precordial-leads | Limb-leads | Precordial-leads | Limb-leads | Precordial-leads | Limb-leads | Precordial-leads |
| PR interval | 1.0% | 0.0% | 0.5% | 0.0% | 0.2% | 0.0% | 0.0% | 0.0% | 4.4% | 0.0% | 1.5% | 0.4% |
| QRS interval | 34.8% | 53.7% | 39.0% | 22.8% | 49.2% | 9.2% | 36.5% | 49.2% | 34.5% | 30.8% | 54.7% | 26.3% |
| ST segment | 0.0% | 10.5% | 0.0% | 7.4% | 0.0% | 0.0% | 0.0% | 0.0% | 0.0% | 8.4% | 0.0% | 1.2% |
| T wave | 0.0% | 0.0% | 0.0% | 30.3% | 7.0% | 34.5% | 0.0% | 14.3% | 0.0% | 21.9% | 0.0% | 16.0% |
| TP interval | 0.0% | 0.0% | 0.0% | 0.0% | 0.0% | 0.0% | 0.0% | 0.0% | 0.0% | 0.0% | 0.0% | 0.0% |
| Total | 35.8% | 64.2% | 39.5% | 60.5% | 56.4% | 43.6% | 36.5% | 63.5% | 38.9% | 61.1% | 56.2% | 43.8% |

## Discussion

The eLVMass-Net was trained on CT-derived LVM values, 12-lead ECG, and demographic information of around 1,500 individual patients. Our study showed that this ECG-grouping-based and demographic-inclusive model outperforms other state-of-the-art deep learning models for LVM estimation. The addition of scalar ECG features such as QRS duration and axis provided insignificant improvement for model performance. Additionally, the sex-specific eLVMass-Net model showed tendency towards better prediction performance than the non-sex-specific model. The alterations in both QRS and T wave voltages associated with increasing LVM may be disparate between both genders.

Our proposed method is effective in predicting LVM values using ECG signals and demographic data as inputs. In the case of the LVM-AI model, the observation of overfitting during training suggests that the model may be too complex or not regularized enough for the size of the dataset used in training. It means that the model has learned to fit the training data very well but needs to generalize better to new data. Compared with the performance between our proposed method and LVM-AI, ours improved by 27% for MAE and MAPE. On the other hand, the ECG segmentation labels are necessary for the ecgAI training pipeline, which is not available in the original XML files. When applying ecgAI on other datasets, additional effort is needed to label ECG segments or the model for the segmentation task. The LVM estimation task will be trained on separate datasets. The experimental results suggest that using a separate dataset for training the ECG segmentation model may have contributed to the low performance of the ecgAI model on our dataset. It may be due to differences in data distribution, recording devices, or preprocessing steps between the two datasets. Therefore, our proposed method shows a relative improvement of 33% for MAE and MAPE compared to ecgAI.

The results of this study are to be further interpreted in the clinical context. First, our proposed model focuses more on the QRS interval of limb leads and T wave in precordial leads with increasing LVM. It is proposed that a hypertrophied heart grows disproportionately towards the inferior, leftward, and posterior axes.^23,24^ Also, T-wave abnormalities may reflect the severity of left ventricular hypertrophy. Respectively integrating both precordial-and limb-lead features by individual encoders may further increase the diagnostic accuracy.^25,26^ Second, previous studies indicated that sex difference exists in QRS duration and voltage regardless of baseline body size or left ventricular mass.^16,27^ Even with similar comorbidities or disease severity, there are significant differences in terms of left ventricular mass and extent of myocardial fibrosis between sexes.^28,29^ The sex-specific model revealed notable improvement in terms of MAPE for predicting LVM compared with the non-sex-specific model. There were distinct differences in segment-wise importance associated with increasing LVM between men and women. Likewise, it was demonstrated that the presentation of either T wave inversions in men or increased precordial voltage in women is associated with heart failure hospitalization.^30^ It is possible that currently developed deep learning algorithms are able to detect important sex-specific pathophysiological differences.^31^

Despite the promising results and contributions of this study, the notable limitation is the absence of external validation data. Although we conducted rigorous experiments and evaluations using carefully curated datasets, the lack of external validation hinders the generalizability of our findings to different populations or datasets. External validation data would provide a valuable opportunity to assess the performance and robustness of our proposed method on unseen and diverse datasets, ensuring its applicability in real-world scenarios. Future research should focus on obtaining and incorporating external validation data to further validate and enhance the reliability and generalizability of our approach.

## Conclusions

Accurate assessment of LVM is crucial in diagnosing and managing cardiovascular diseases. We proposed eLVMass-Net as a novel approach that includes relevant heartbeat waveforms, inter-lead grouping, and demographic information for LVM estimation. Our model architecture incorporates pre-processing techniques that focus on synchronized heartbeat waveforms and ECG groups based on different projection planes to improve the understanding of their relationships. For sex disparities, the sex-specific model is able to discriminate important ECG features associated with left ventricular mass.

## Abbreviations

CNN: convoluted neuron network
LVH: left ventricular hypertrophy
LVM: left ventricular myocardial mass
MAE: mean absolute error
MAPE: mean absolute percentage error
ML: machine learning
MLP: multilayer perceptron
SOTA: state of the art
TCN: temporal convoluted network

## Data Availability

National Taiwan University Hospital has the proprietary right to all patient data involved in the study, which was derived from the Taiwan CVAI dataset.

